# A Systematic Review of the Association between Sedentary Behavior and Non-Motor Symptoms of Parkinson’s Disease

**DOI:** 10.1101/2023.10.12.23296941

**Authors:** Aiza Khan, Joy Ezeugwa, Victor Ezeugwu

## Abstract

**Background:** Parkinson’s disease, known for motor symptoms, often brings early non-motor issues that severely affect patients’ quality of life. While there are not many effective treatments, physical activity and exercise can help. Yet, another component of the movement intensity continuum warrants examination—prolonged sitting or sedentary behavior. Thus, aim of this study was to perform a systematic review to investigate the relationship between sedentary behaviour and non-motor symptoms, specifically cognitive decline, depression and sleep deficits in Parkinson’s disease.

**Methods:** Conforming to PRISMA guidelines, we reviewed the literature up to February 28, 2023, to investigate how sedentary behavior correlates with non-motor symptoms such as cognitive impairment, depression, and sleep disturbances in Parkinson’s disease. A systematic search of the literature was conducted via electronic databases including MEDLINE, CINAHL, Scopus, PubMed and PsycINFO. The eligibility criteria for study selection was: (i) If it studied the Individuals with Parkinson’s disease with sedentary behaviour(iii) studies investigating the association between sedentary behavior and at least one of the non-motor symptoms, including depression, sleep quality, and/or cognitive impairment. New-castle-Ottawa scale for cross-sectional and cohort studies was used to perform quality assessment of the studies.

**Results:** Of the 463 publications found, 7 studies met the inclusion criteria. All the studies were observational. Total number of cases across all studies were 980. Collectively, these studies show that prolonged sedentary time is associated with increased cognitive, depressive, and sleep-related problems.

**Conclusion:** Recognizing sedentary behavior as an independent factor holds pivotal significance. The intricate relationship between sedentary behavior and Parkinson’s disease non-motor symptoms necessitates further exploration to potentially enhance therapeutic strategies for those living with the disease.

## 1. Introduction

1.1 Parkinson’s disease (PD) is a progressive neurodegenerative disorder affecting 1 to 2% of adults over age 65 years and 4% of adults over age 80 years (1). PD has been identified as the “fastest growing neurological disorder” between 1990 and 2016, contributing to a significant number of deaths and disabilities globally (2). In 2019, PD was attributed with 5.8 million disability-adjusted life-years (DALYs) across 195 countries, representing an 81% increase since 2000 (2). Click or tap here to enter text.Clinically, PD is primarily characterized by motor symptoms such as bradykinesia, resting tremor, rigidity, gait abnormalities, and postural impairment (1). However, it is important to note that a diverse range of non-motor symptoms (NMS) also play a significant role in PD symptomology (3)(4). These NMS include sleep disturbances, sensory deficits, mood disorders including depression, apathy, or anxiety, autonomic nervous system dysfunction (orthostatic hypotension and obstipation), olfactory dysfunction, and cognitive impairment (3)(4). Several studies indicate that NMS often precede motor symptoms and can have a more profound impact on the quality of life of individuals living with PD(5). Mood disorders such as depression and anxiety, as well as pain, fatigue, and cognitive impairment contribute to the disease burden as PD progresses (3)(6).

Despite their significance, NMS frequently go unrecognized and undertreated, posing a challenge for people living with PD (7). Moreover, NMS in PD may present significant challenges that affect day-to-day functioning and physical activity levels, as many people with PD often struggle to meet the recommended guidelines of 150 minutes per week of moderate-intensity physical activity(8). In other words, activities that are intense enough to breathe heavily and break a sweat. With movement challenges and NMS experienced by people living with PD, moving fast or long enough to reach recommended targets may be hard. As a consequence, people with PD often lead sedentary lifestylesClick or tap here to enter text.(9)(10), which can have adverse short-and long-term effects on their health (11).

1.2.“Sedentary behavior, characterized by activities in sitting or lying with minimal energy expenditure (12), is more prevalent in individuals with PD compared to age-matched adults (13). For example, people with PD engage in approximately 10 hours of sedentary behaviors during waking hours, often in longer bouts compared to their age-matched peers without PD (13). Notably, sedentary behavior is a well-established risk factor for chronic diseases such as diabetes, cancer, cardiovascular disease, and negatively affects mental health conditions like stress, anxiety, depression, and dementia (14). Understanding the potential association between sedentary behavior and non-motor symptoms in PD is crucial for identifying underlying mechanisms and improving symptom management (3)(13). Previous studies have shown positive effects of exercise on both motor and non-motor symptoms in PD(15), however exercise and sedentary behaviour are distinct and independent factors (13)(16). Although a potential link between sedentary behaviour and cognitive impairment has been suggested in PD (13), clear associations between non-motor symptoms and sedentary behavior are yet to be established, but determining such links may help in better management of NMS. As the number of PD cases continues to rise globally, understanding and managing NMS are essential for improving the quality of life of individuals with PD (7). Understanding the distinctive associations between sedentary behavior and NMS could inform future interventions for PD (17)(18) (16).

In this paper, authors aim to review the literature for studies that highlight the associations between sedentary behavior (other search terms may include sitting/lying or physical inactivity) with non-motor symptoms in individuals living with PD. More specifically, authors will focus on the associations between non-motor symptoms related to mental health such as sleep quality, depression, and cognitive impairments with sedentary behavior in individuals living with PD.

## 2. Methods

### 2.1 Protocol and registration

In order to report this Systematic review, we used Preferred Reporting Items for Systematic Reviews and Meta-Analyses (PRISMA) statement (19)as shown in S1 Table. The protocol was registered with PROSPERO (ID: CRD42023405422).

### 2.2 Identification of relevant studies

A systematic search was conducted to identify all original peer-reviewed articles available on the association of sedentary behavior with non-motor symptoms, including sleep, depression/anxiety, and cognitive decline in Parkinson’s disease until February 28, 2023. The search was performed on several databases, including MEDLINE, CINAHL, Scopus, and PubMed and PsycINFO using different combinations of search terms such as non-motor symptom* OR depress* OR anxiety OR mood AND/OR sleep* OR insomnia* OR sleep difficulties OR reduced sleep AND/OR cognitive decline, OR cognitive impairment OR sedentary AND sitting OR bed-ridden OR lying down OR physical inactivity OR lack OR minimal physical-activity, in different combinations with Parkinson’s disease.

Next, all identified studies were transferred to Covidence software for screening.

The eligibility criteria were based on the PI(E)COS (participants, intervention (exposure), context, outcomes, and study design) framework(20). The original peer-reviewed articles were included if they met specific criteria. First, studies were selected if they included participants with PD who exhibited non-motor symptoms at the time of inclusion in the study. Second, the exposure of interest was sedentary behavior, as the primary focus of our study was to identify the association between non-motor symptoms and sedentary behavior. Regarding context/comparators, the comparator criterion was not applicable to our study. Patients from all settings, such as the community or hospitalized, were considered for inclusion in the study. In terms of outcomes, all the included studies focused on the association between sedentary behavior and at least one of the non-motor symptoms, including depression, sleep quality, and/or cognitive impairment. Finally, all types of study designs were included as this is a relatively new area of study with limited available data. Therefore, all peer-reviewed articles written in English describing original quantitative research were included. Additionally, the reference lists of all eligible studies were carefully examined to identify any additional relevant studies.

### 2.3 Selection of relevant studies

Based on the eligibility criteria mentioned above, two independent reviewers screened the articles for selection. At first, the title and abstract screening was performed which was followed by the full-text screening. All conflicts between the two reviewers were discussed and resolved by the third reviewer and a consensus was reached.

### 2.4 Data Synthesis

The data from the selected studies were extracted and organized using an Excel spreadsheet and Covidence software. The following information was collected: study number, title of study, first author, objective(s) of study, study design, year, country, number of participants, exposure setting, age, sex/gender, type of non-motor symptoms, type of test for non-motor symptom, measurement of sedentary behavior, main outcomes, secondary outcomes, key findings, and statistical analysis (available as supplementary material).

Due to factors such as small number of studies, variation in study designs of studies and methods, and heterogeneity, it was determined that a meta-analysis may not yield worthwhile results. Therefore, a narrative synthesis was conducted, following Popay’ et al.’s instructions ad suggestions (21) A tabulation format was chosen to describe the characteristics of included studies, the characteristics of participants, and the objectives and outcomes related to the association between sedentary behavior and non-motor symptoms. The protocol for the systematic review was registered on PROSPERO (ID: CRD42023405422) on April 11, 2023.

### 2.5 Quality assessment

For the longitudinal studies Newcastle Ottawa scale for cohort studies was used. For cross sectional studies, quality Appraisal was performed studies Newcastle-Ottawa Scale adapted for cross-sectional studies. Studies scoring 5 or more were considered to be good quality studies(22,23).

## 3 Results

Out of 463 articles, 183 duplicate articles were removed, leaving 280 studies. Of these, 243 were considered irrelevant, and 37 studies underwent full-text screening. Finally, 7 studies(13)(24)(25)(26)(27)(28)(29) were found eligible for systematic review and included in the data extraction and narrative synthesis. The selection process is summarized in a PRISMA flowchart (Figure 1).

**Figure 1.**
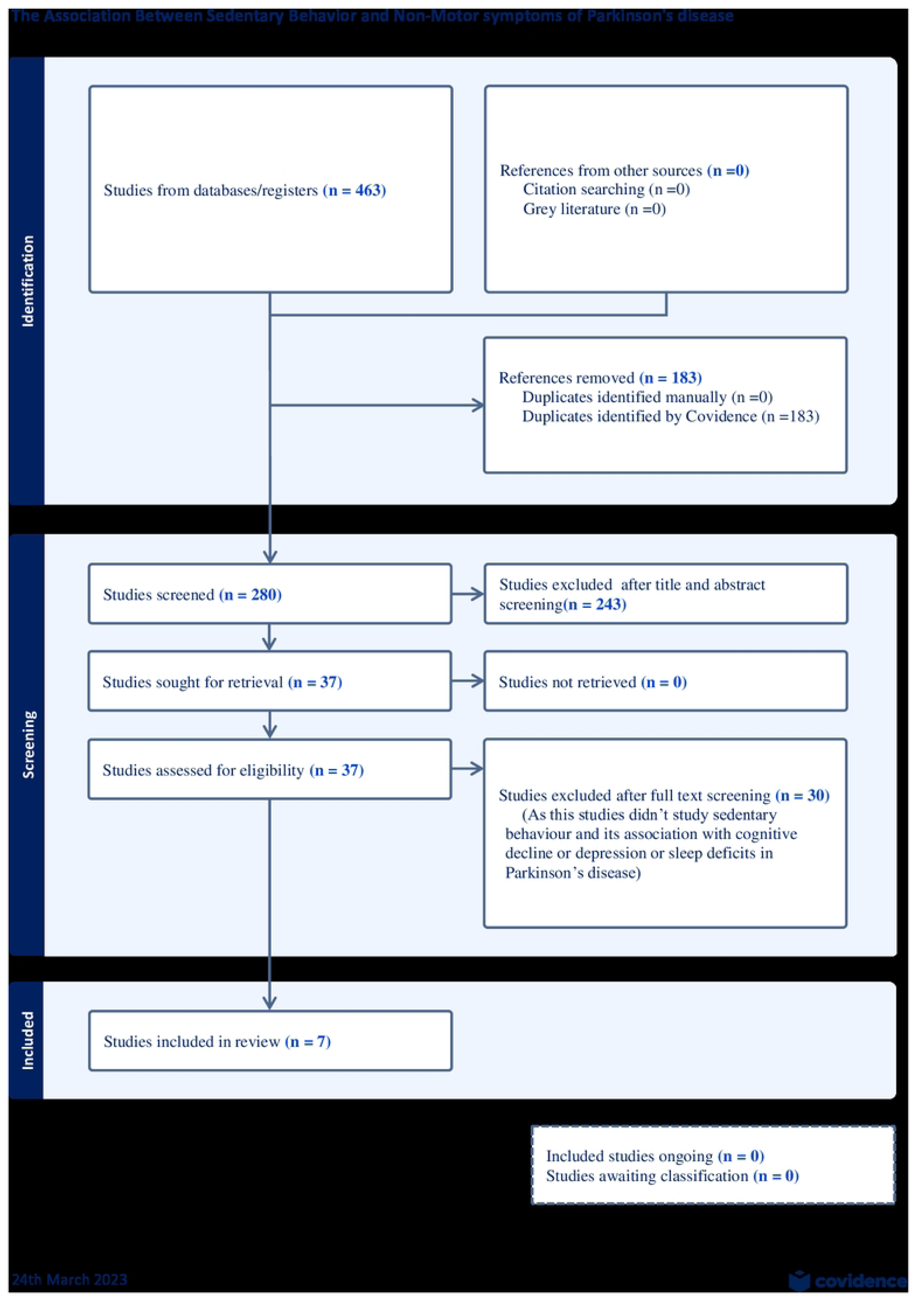
Depicting the PRISMA Steps followed to select articles for the current study.

Table 1 shows the characteristics of the included studies, while Table 2 presents the characteristics of people with PD. All 7 studies selected were observational studies, with three being cross-sectional and three being secondary analyses of longitudinal cohort studies. While one of them was exploratory observational study. Five studies were conducted in the USA, and the remaining two were done in the United Kingdom. Total number of cases across all studies were 980 (excluding controls). Notably, all the studies were conducted within the last decade, reflecting the emerging trend in research to better understand the relationship between sedentary behavior and non-motor symptoms in PD. Six studies assessed the correlation of sedentary behavior and cognition, with two studies including an assessment of depression in addition to cognition. Only one study studied the link between sedentary behavior and sleep. The studies were primarily carried out in community settings, except for one that was conducted in a home-based setting.

**Table 1.**
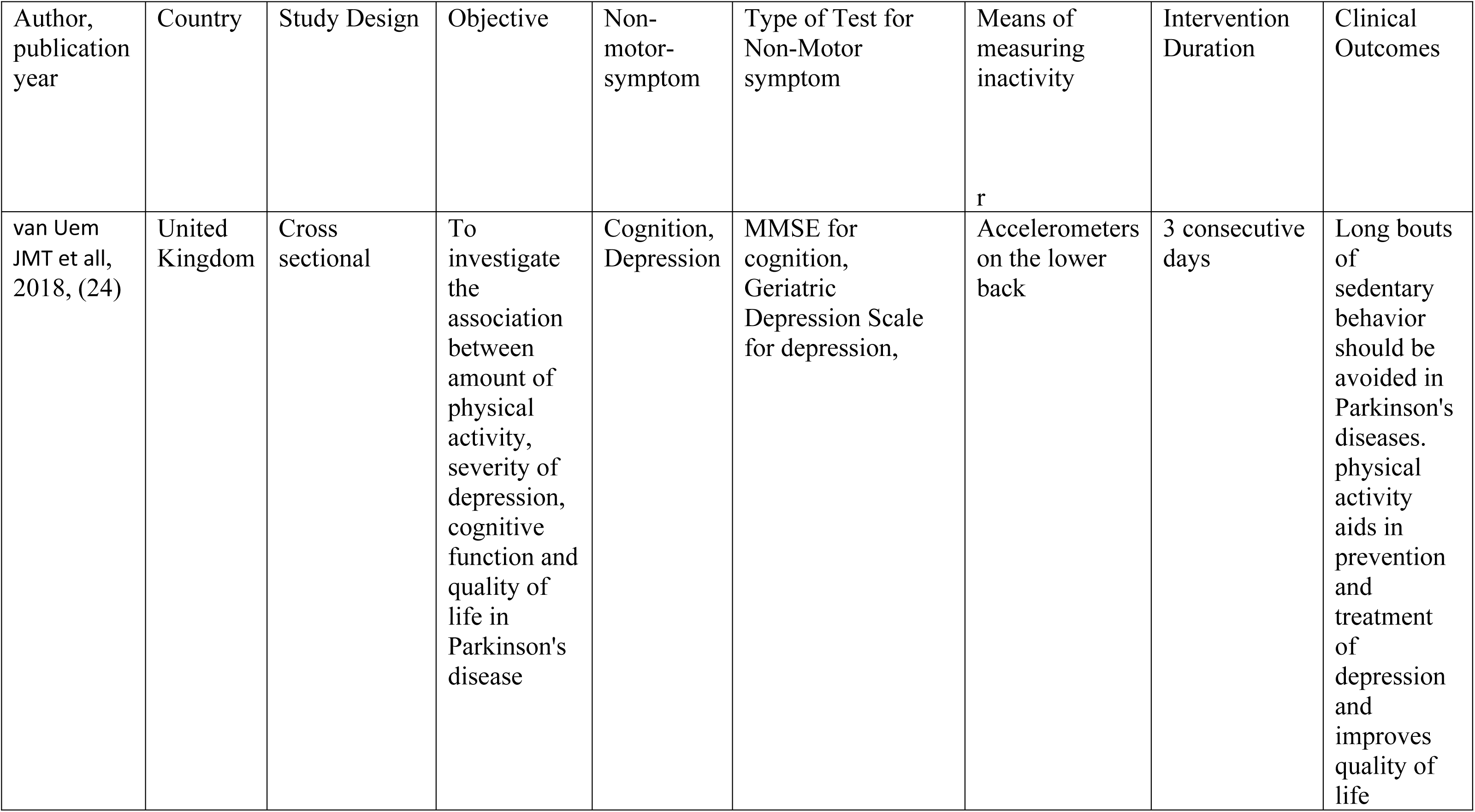
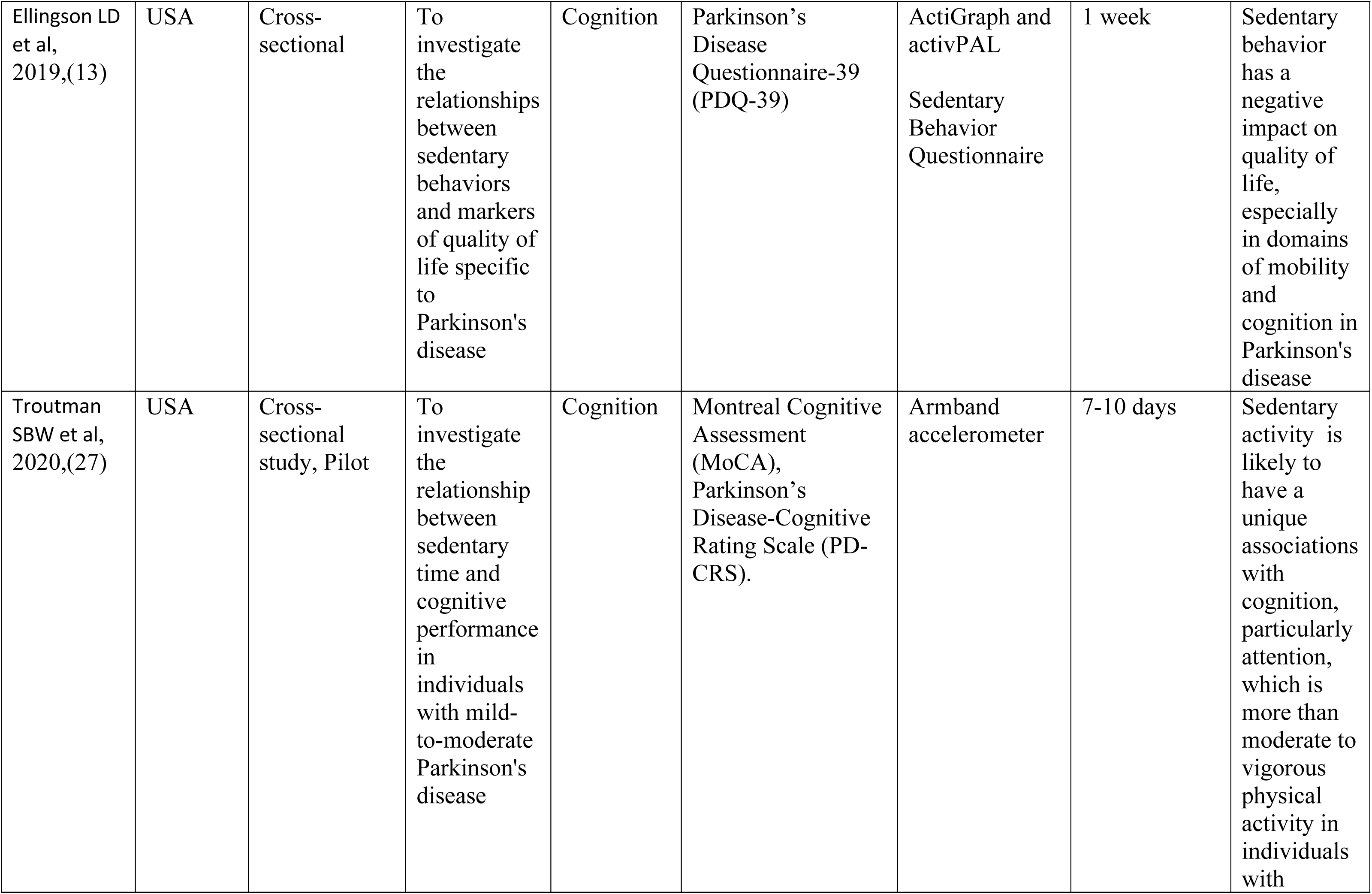
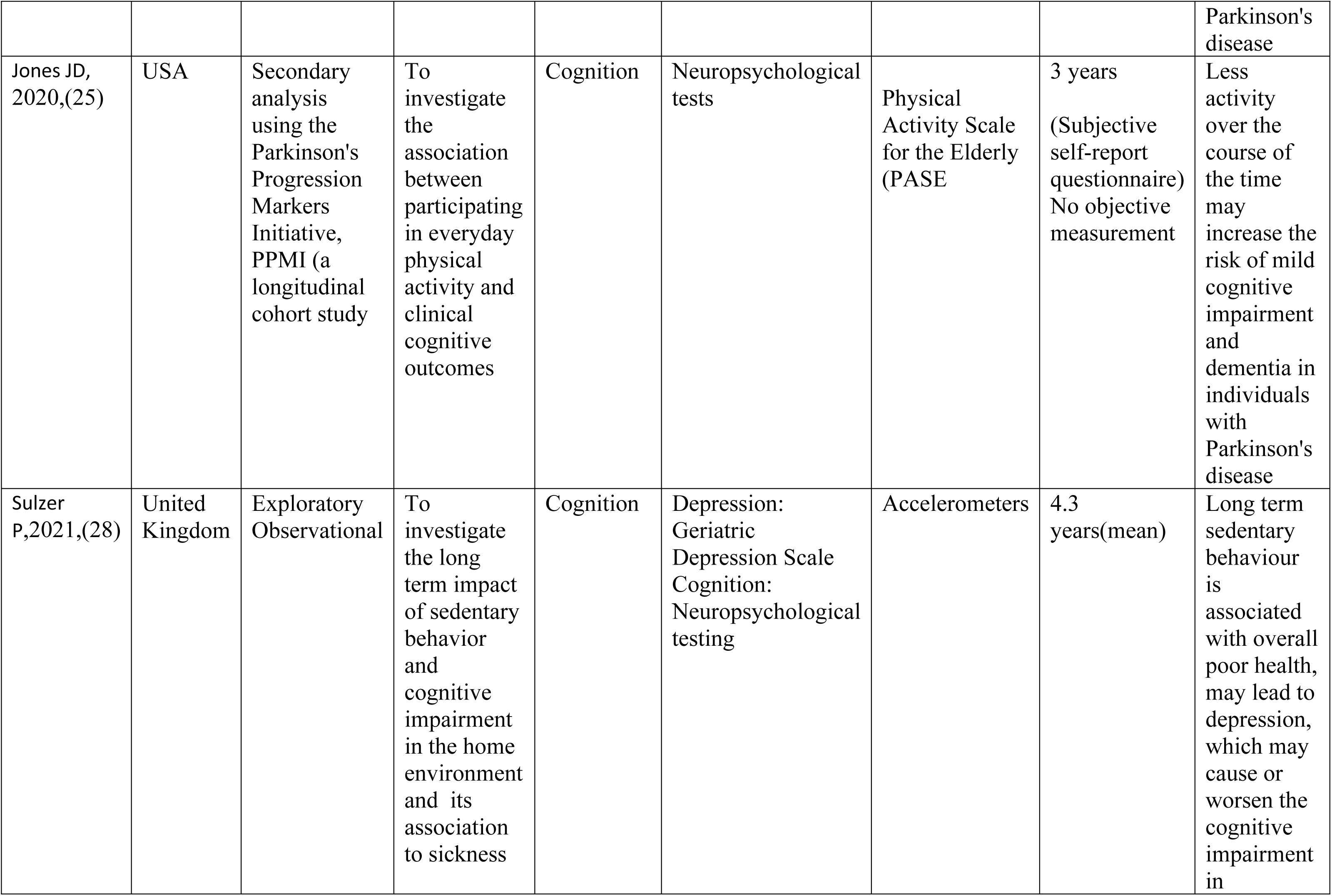
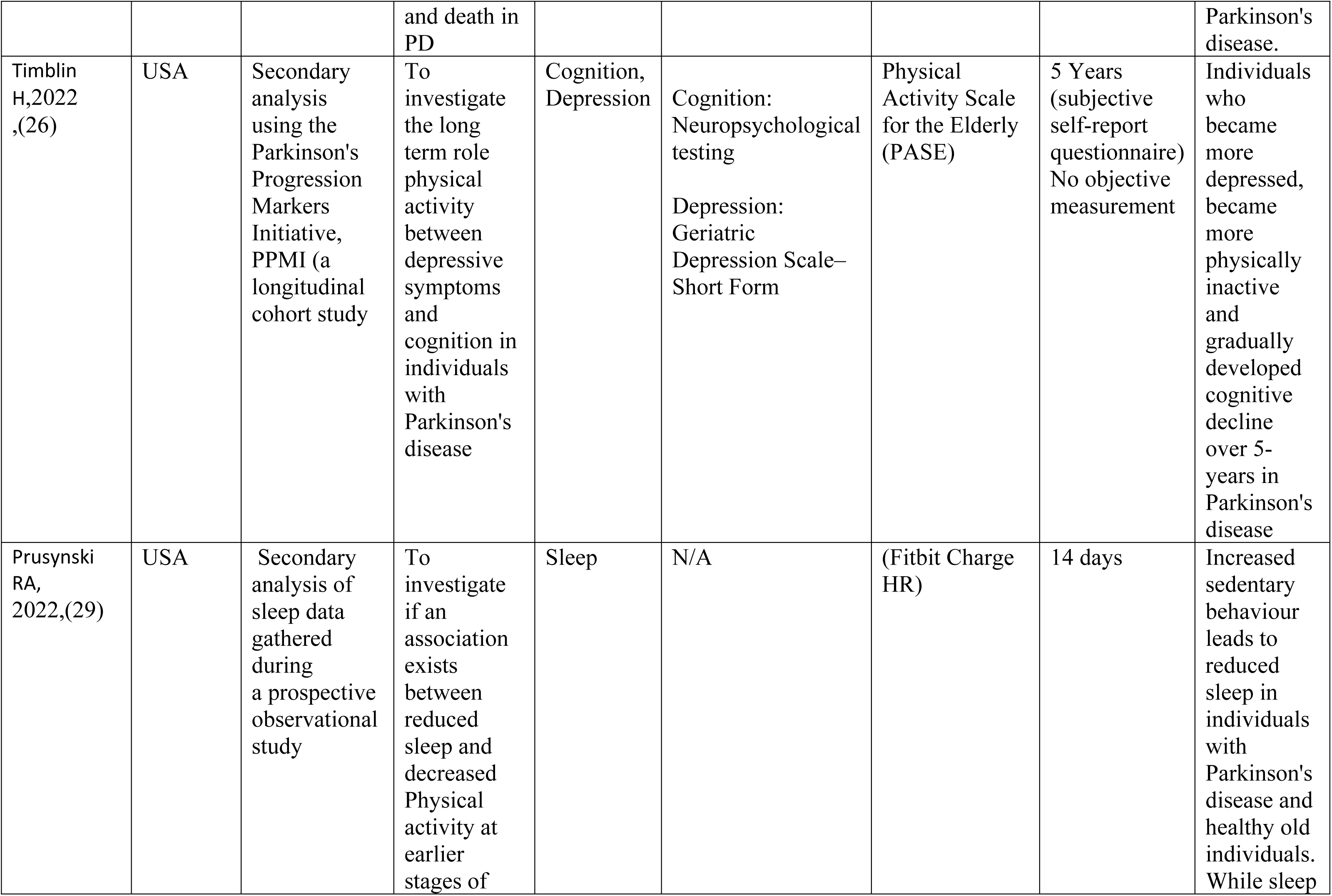
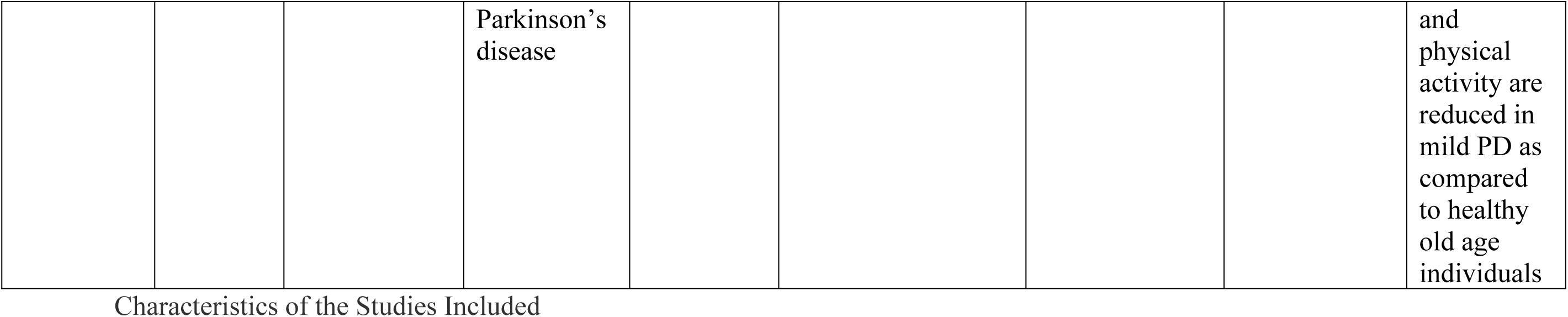

| Author,<br>publication<br>year | Country | Study Design | Objective | Non-motor-<br>symptom | Type of Test for<br>Non-Motor<br>symptom | Means of<br>measuring<br>inactivity<br>r | Intervention<br>Duration | Clinical<br>Outcomes |
| --- | --- | --- | --- | --- | --- | --- | --- | --- |
| van Uem<br>JMT et al,<br>2018, (24) | United<br>Kingdom | Cross<br>sectional | To<br>investigate<br>the<br>association<br>between<br>amount of<br>physical<br>activity,<br>severity of<br>depression,<br>cognitive<br>function and<br>quality of<br>life in<br>Parkinson's<br>disease | Cognition,<br>Depression | MMSE for<br>cognition,<br>Geriatric<br>Depression Scale<br>for depression, | Accelerometers<br>on the lower<br>back | 3 consecutive<br>days | Long bouts<br>of<br>sedentary<br>behavior<br>should be<br>avoided in<br>Parkinson's<br>diseases.<br>physical<br>activity<br>aids in<br>prevention<br>and<br>treatment<br>of<br>depression<br>and<br>improves<br>quality of<br>life |
| Ellingson LD et al, 2019,(13) | USA | Cross-sectional | To investigate the relationships between sedentary behaviors and markers of quality of life specific to Parkinson's disease | Cognition | Parkinson's Disease Questionnaire-39 (PDQ-39) | ActiGraph and activPAL<br>Sedentary Behavior Questionnaire | 1 week | Sedentary behavior has a negative impact on quality of life, especially in domains of mobility and cognition in Parkinson's disease |
| Troutman SBW et al, 2020,(27) | USA | Cross-sectional study, Pilot | To investigate the relationship between sedentary time and cognitive performance in individuals with mild-to-moderate Parkinson's disease | Cognition | Montreal Cognitive Assessment (MoCA), Parkinson's Disease-Cognitive Rating Scale (PD-CRS). | Armband accelerometer | 7-10 days | Sedentary activity is likely to have a unique associations with cognition, particularly attention, which is more than moderate to vigorous physical activity in individuals with |
|  |  |  |  |  |  |  |  | Parkinson's disease |
| Jones JD, 2020,(25) | USA | Secondary analysis using the Parkinson's Progression Markers Initiative, PPMI (a longitudinal cohort study) | To investigate the association between participating in everyday physical activity and clinical cognitive outcomes | Cognition | Neuropsychological tests | Physical Activity Scale for the Elderly (PASE | 3 years<br>(Subjective self-report questionnaire)<br>No objective measurement | Less activity over the course of the time may increase the risk of mild cognitive impairment and dementia in individuals with Parkinson's disease |
| Sulzer P,2021,(28) | United Kingdom | Exploratory Observational | To investigate the long term impact of sedentary behavior and cognitive impairment in the home environment and its association to sickness | Cognition | Depression: Geriatric Depression Scale<br>Cognition: Neuropsychological testing | Accelerometers | 4.3 years(mean) | Long term sedentary behaviour is associated with overall poor health, may lead to depression, which may cause or worsen the cognitive impairment in |
|  |  |  | and death in PD |  |  |  |  | Parkinson's disease. |
| Timblin H,2022 ,(26) | USA | Secondary analysis using the Parkinson's Progression Markers Initiative, PPMI (a longitudinal cohort study | To investigate the long term role physical activity between depressive symptoms and cognition in individuals with Parkinson's disease | Cognition, Depression | Cognition: Neuropsychological testing<br>Depression: Geriatric Depression Scale– Short Form | Physical Activity Scale for the Elderly (PASE) | 5 Years (subjective self-report questionnaire)<br>No objective measurement | Individuals who became more depressed, became more physically inactive and gradually developed cognitive decline over 5-years in Parkinson's disease |
| Prusynski RA, 2022,(29) | USA | Secondary analysis of sleep data gathered during a prospective observational study | To investigate if an association exists between reduced sleep and decreased Physical activity at earlier stages of | Sleep | N/A | (Fitbit Charge HR) | 14 days | Increased sedentary behaviour leads to reduced sleep in individuals with Parkinson's disease and healthy old individuals. While sleep |
|  |  |  | Parkinson's disease |  |  |  |  | and physical activity are reduced in mild PD as compared to healthy old age individuals |

Characteristics of the Studies Included

**Table 2.**

| Author<br>,<br>publica<br>tion<br>year | Non motor<br>symptom<br>studied | Number of<br>Participants<br>(Intervention<br>N) | Setting | Age of<br>patients | Biological sex<br>(Male/Female<br>%) |
| --- | --- | --- | --- | --- | --- |
| van<br>Uem<br>JMT et<br>all,<br>2018,<br>(24) | Depression,<br>Cognition | 47 | Home | 65-74 (mean<br>age 70) | Male 74%, 26%<br>female |
| Ellingso<br>n LD et<br>al, | Cognition | 52 | Community | 59.9-75.7(mean<br>age:67.8) | 56% were male,<br>44% female |
| 2019,(<br>13) |  |  |  |  |  |
| Troutman SBW<br>et al,<br>2020,(<br>27) | Cognition | 17 | Community | 50-80 (mean<br>age:65.07) | 82% male, 18%<br>female. |
| Jones<br>JD,<br>2020,(<br>25) | Cognition | 307 | Community | 40-80 (mean<br>age:61) | 65.8<br>%male,34.2%<br>female |
| Sulzer<br>P,2021,<br>(28) | Cognition | 45 study<br>completed:20. | Community | 44-80(mean<br>age: 67.5) | (78%) male 12%<br>females |
| Timblin<br>H,2022<br>,(26) | Depression | 487 | Community | 33-84 (mean<br>age: 61) | 65.1% male,<br>34.1 % females |
| Prusynski RA, 2022,(29) | Sleep | 25 | Community | 69 (mean age) | not specified |

Characteristics of the Patients Included in the study.

The studies were further categorized into three groups based on the non-motor symptoms studied as follows:

### 3.1 Sedentary Behaviour and Cognitive Changes in Parkinson’s Disease

Out of the seven studies, six investigated the impact of sedentary behavior on cognition (13)(27)(26)(24)(25)(28). Three of these studies focused exclusively on cognition(28)(27)(25), while the other three assessed depression and other factors in addition to cognition(13)(26)(24). All the studies were observational studies encompassing different study designs. It was consistently reported by all six studies that a sedentary lifestyle was associated with cognitive decline in PD(13)(27)(Timblin et al., 2022) (Timblin et al., 2022)(24)(25)(28). Interesting variations were observed in the aspects of this relationship investigated by the different studies. One study found an association between sedentary behavior and an elevated risk of developing PD-related mild cognitive impairment and dementia(25). Another study noted that as leisure/recreational physical activities declined over time, the risk of developing PD-related dementia increased (25).

Similarly, another study reported that sedentary behavior in individuals with PD resulted in overall poor health, particularly worse in those with cognitive decline (28). A study by Troutman et al. reported that sedentary activity negatively affected cognition in general, particularly attention, and also led to reduced cognitive processing and communication, resulting in a poorer quality of life (27). These studies highlighted the cumulative detrimental effects of sedentary behavior and physical inactivity on cognition in PD, subsequently lowering the quality of life of people with PD (13)(24).

### 3.2 Sedentary Behavior and Depression/Anxiety in Parkinson’s disease

Two studies assessed depression in addition to the cognitive impairment in individuals with PD(26)(24). One study reported that depression led to sedentary behavior, which in turn, led to cognitive decline over time(26). This suggests that lack of physical activity might play a role in depression and its eventual impact on cognitive decline(26). Similarly, another study determined that PD patients benefit from being physically active, while long periods of sedentary behaviour may contribute to the etiology of depression(24). Being physically active not only improved quality of life but may also helped in the prevention and therapy for depression in PD (24).

### 3.3 Sedentary behavior and sleep deficits in Parkinson’s disease

Only one study assessed the association between sleep and sedentary behaviour in PD(29). The study reported that individuals with mild PD slept less and were less active compared to a group of healthy older adults. Of note, in both groups, less sleep was related to more sedentary behaviour (29).

### 3.4 Factors contributing to Sedentary behaviour in Parkinson’s disease

It is well-established that motor symptoms typically lead to physical inactivity in PD(30). However, it is crucial to highlight here that lack of physical activity is associated with cognitive impairment independent of the severity of motor symptoms (25). One study reported an association between sedentary behavior and cognitive decline in patients newly diagnosed with PD, when motor symptoms are less severe (25). These findings make it difficult to establish causation. Moreover, it has been reported that mood and thought disorders such as apathy and anxiety may increase the risk of physical inactivity (26). Additionally, factors such as female gender, older age, and a decline in overall physical capacity are important determinants of sedentary behavior (31).

### 3.5 Quality Appraisal

Using the Newcastle-Ottawa Scale, six out of the seven studies received a rating of good quality (score > 5), while one study was rated as moderate quality (score < 5), as illustrated in Figures 2a and 2b. Among the six studies rated as good quality, three were cross-sectional, while the remaining three were longitudinal studies.

**Figure 2a.**
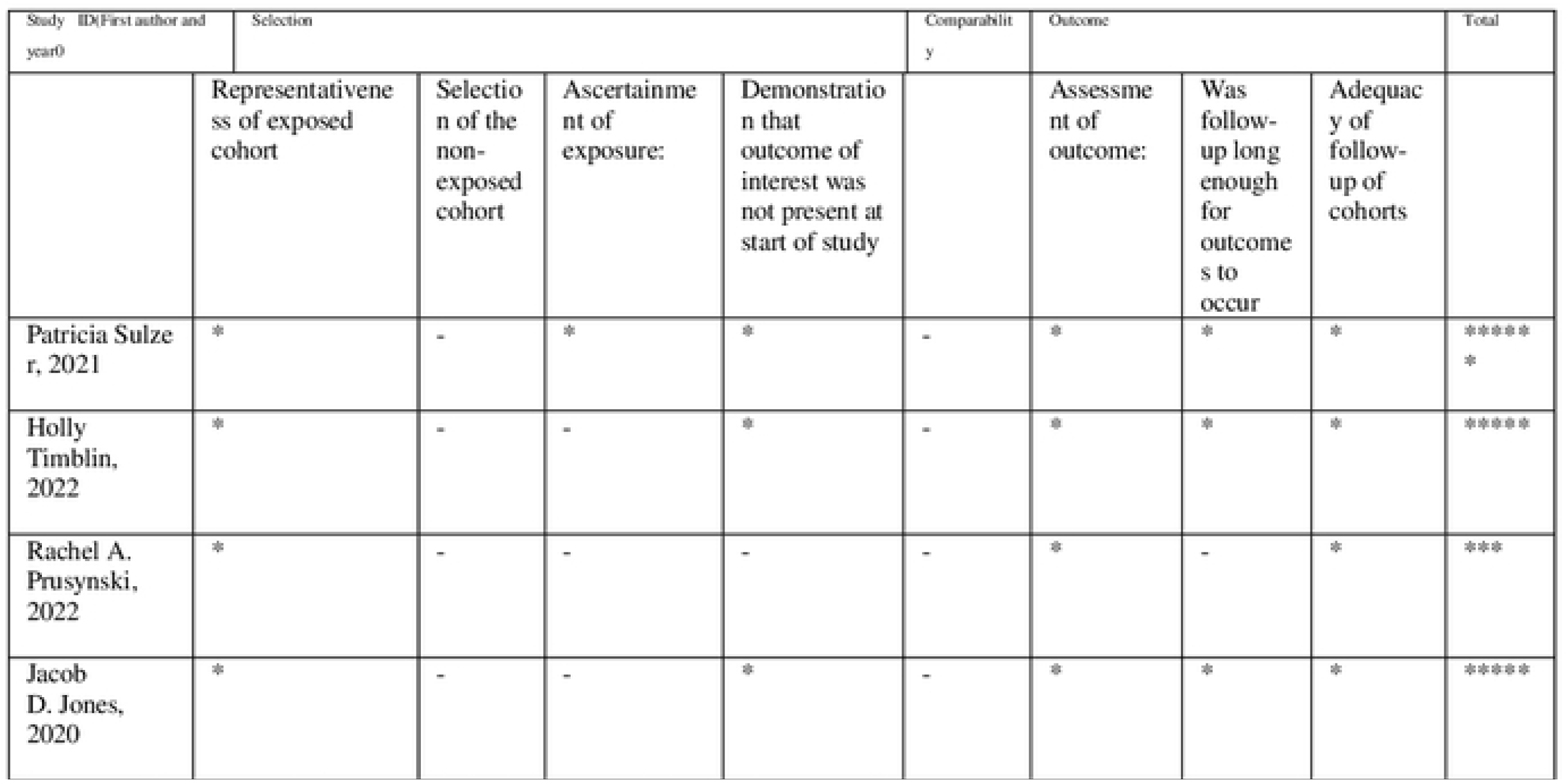
Quality Appraisal for longitudinal studies using New Castle Ottawa.

***Figure 2b.**
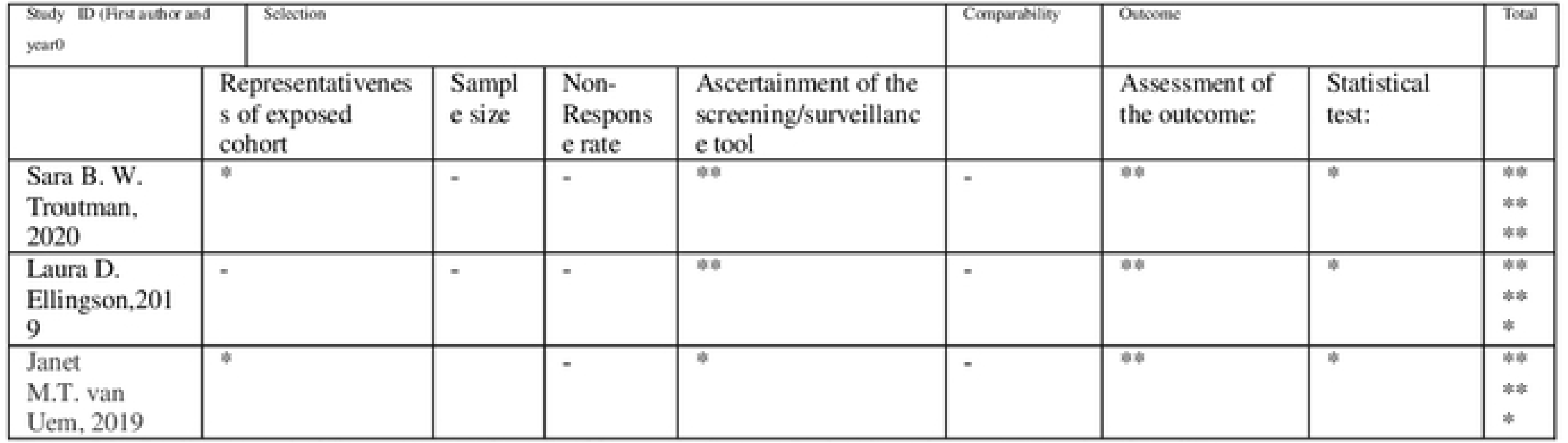
Quality Appraisal for case-control studies Newcastle-Ottawa Scale adapted for cross-sectional studies.

Scale:

Study acquiring at least 5 stars, or more is considered good quality. Three main categories Selection, Comparability and Outcomes with further subdivisions.

Selection 1) Representativeness of the exposed cohort a) Truly representative (one star) b) Somewhat representative (one star) c) <u>No star for</u>: elected group d) No description of the derivation of the cohort 2) Selection of the non-exposed cohort a) Drawn from the same community as the exposed cohort (one star) <u>No star for:</u> b) Drawn from a different source c) No description of the derivation of the non exposed cohort 3) Ascertainment of exposure a) Secure record (e.g., surgical record) (one star) b) Structured interview (one star) No star for: c) Written self report d) No description e) Other 4) Demonstration that outcome of interest was not present at start of study a) Yes (one star) <u>No star if:</u> b) No Comparability 1) Comparability of cohorts on the basis of the design or analysis controlled for confounders a) The study controls for age, sex and marital status (one star) b) Study controls for other factors (list) (one star) <u>No star if</u>: c) Cohorts are not comparable on the basis of the design or analysis controlled for confounders Outcome 1) Assessment of outcome a) Independent blind assessment (one star) b) Record linkage (one star) c) <u>No star for:</u> Self report d) No description e) Other 2) Was follow-up long enough for outcomes to occur a) Yes (one star) b) No Indicate the median duration of follow-up and a brief rationale for the assessment above: 3) Adequacy of follow-up of cohorts a) Complete follow up-all subject accounted for (one star) b) Subjects lost to follow up unlikely to introduce bias-number lost less than or equal to 20% or description of those lost suggested no different from those followed. (One star) <u>No star for:</u> c) Follow up rate less than 80% and no description of those lost d) No statement.

Study acquiring at least 5 stars, or more is considered good quality. Three main categories Selection, Comparability and Outcomes with further subdivisions.

Selection: (Maximum 5 scores) 1) Representativeness of the cases: a) Truly representative of the HCC patients (consecutive or random sampling of cases). 1 star b) Somewhat representative of the average in the HCC patients (non-random sampling). 1 star. No star for: c) Selected demographic group of users. d) No description of the sampling strategy. 2) Sample size: a) Justified and satisfactory (≥ 400 HCC included). 1 star. No star for: b) Not justified (<400 HCC patients included). 3) Non-Response rate a) The response rate is satisfactory (≥95%). 1 Star. No star if: b) The response rate is unsatisfactory (<95%), or no description. 4) Ascertainment of the screening/surveillance tool: a) Validated screening/surveillance tool. 2-star b) non-validated screening/surveillance tool, but the tool is available or described. 1 star. No star if: No description of the measurement tool. Comparability: (Maximum 1 stars) 1) The potential confounders were investigated by subgroup analysis or multivariable analysis. a) The study investigates potential confounders. 1 tar. No star if: b) The study does not investigate potential confounders. Outcome: (Maximum 3 stars) 1) Assessment of the outcome: a) Independent blind assessment. 2 stars b) Record linkage. 2 stars c) Self report. 1 Star. No star if d) No description. 2) Statistical test: a) The statistical test used to analyze the data is clearly described and appropriate. 1 star. No star if: b) The statistical test is not appropriate, not described or incomplete.

*: star, -: no star

: star, -: no star

## 4. Discussion

There is substantial evidence that physical activity is highly beneficial for individuals with PD, potentially delaying or slowing down the disease progression (32)(33). Exercise has also been demonstrated to improve non-motor symptoms of PD (34). However, it is important to recognize that sedentary behavior is not merely the opposite of physical activity or exercise, but an independent behavior that is associated with adverse health outcomes(13)(14). The unique relationship between sedentary behavior and non-motor symptoms of PD has been suggested by some researchers(13)(35). Yet, a clearer understanding of this relationship is required, considering that the prevalence of PD is increasing, and technological advances may encourage even more sedentary behaviors. Although in recent years, this gap has been highlighted(36). It has been suggested that sedentary behavior and its role in non-motor symptoms is uniquely essential (13). Hence in the context of PD, further understanding of sedentary behavior and its objective measurements would provide another additional dimension, further understanding of which is crucial to determine further gaps that can be filled, subsequently enabling researchers to provide enhanced therapeutic strategies for individuals living with PD.

In this study, we specifically examined the association between sedentary behavior and three non-motor symptoms namely cognitive impairment, depression and sleep in the context of PD. A brief discussion on each one of them follows.

### 4.1 Association of Sedentary behavior and Cognition in Parkinson’s disease

Cognitive impairment is a common, non-motor symptom of PD, with a wide spectrum of symptoms related to attention, working memory, visuospatial, and executive functioning (37)(38). The symptoms related to impaired cognition may include worsening of attention, problems with, difficulties with (37)(38). The underlying mechanisms contributing to cognitive decline in PD are complex and may involve genetic factors such as α-synuclein toxicity, Lewy body accumulation, synaptic changes, inflammatory process, and neurotransmitter disruptions (17)(37)(38)(39). Pharmacological interventions are typically used to target these changes in order to enhance cognition, however, the overall improvement remains modest(38). In addition to pharmacological strategies, non-pharmacological strategies, such as physical activity, have been shown to improve various biological mechanisms related to cognition in individuals with PD(37)(38). Thus physical activity is a promising approach in improving cognition in PD(40). Importantly, a distinctive and independent association between cognitive impairment in PD and sedentary behavior has also been suggested,(13)(26) however, yet to investigated in depth. We found studies (Table 1) assessing this link and all of them reported a negative impact of sedentary behavior on cognition. Studies have shown that sedentary behavior can be related with poor global cognition, and cognitive decline(26,28). Furthermore, a prolonged sedentary behavior causes a progress in cognitive decline.

### 4.2 Association of Sedentary Behavior and Depression in Parkinson’s disease

Depression is another common non-motor symptom in PD (41). Depression often manifests before the onset of motor symptoms, though it can occur at any stage of PD (41)(42). Approximately 40% of patients with PD experience symptoms of depression (42). These symptoms may include excessive feeling of sadness, helplessness, lack of concentration, loss of interest in previously enjoyed activities, increased exhaustion, irritability, and dysphoria. Diagnosing depression in PD can be challenging due to symptom overlap with PD symptoms, leading to the condition being untreated in some individuals living with PD(41)(42). Depressive disturbances not only cause emotional burdens, but also have a significant negative impact on quality of life(42). Moreover, depression also negatively affects motor symptoms, and worsens cognitive and functional disabilities(24)(42). It is noteworthy that depression along with anxiety are among the non-motor symptoms that are considered to be the most important predictors of quality of life in individuals with PD(24)(43).

The pathophysiology of depression in PD is not fully understood(41)(43). Various studies suggest that depression in PD could be related to dysfunction of multiple brain areas, such as subcortical nuclei as well as prefrontal cortex, striatal–thalamic–prefrontal and bitemporal limbic circuits. Also, degenerative changes in neurotransmitter systems of the brain may play a role(41)(43). Psychosocial factors have also been suggested to contribute to the etiology of depression in PD (41)(43)(42). Another factor less studied attributed to depression in PD is sedentary behavior (33). One study evaluating the role of physical activity in PD showed that patients with prolonged sedentary behavior displayed more apathetic behavior as compared to others who are active (33). Other studies have also demonstrated a link between physical inactivity and an increased likelihood of depression in PD (26)(14)(34). In this review, we found only two studies that assessed the link between depression and sedentary behavior (26)(24). Both studies reported a potential link between depression and sedentary behavior which may also contribute to cognitive decline(26)(24).

### 4.3 Association of Sedentary Behavior and Sleep in Parkinson’s disease

Sleep-related disorders are highly prevalent in PD (44), and constitute an integral part of spectrum of non-motor symptoms of PD (45). These sleep disorders may present earlier in the course of disease (44). The etiology of sleep disturbances in PD is multifactorial and may include PD-related neurodegenerative processes and medication side-effects(46). A diverse array of sleep disorders is associated with PD, ranging from poor sleep quality, fragmented sleep, reduction in total sleep time, and overall sleep and rapid eye movement behavior disorders (3). Sleep disturbances significantly impact the quality of life in individuals with PD and tend to worsen as the disease progresses(46). In addition, sleep problems may also have a negative effect on the severity of motor and other non-motor symptoms, making effective treatment crucial (3)(46). Various therapeutic interventions are being investigated to improve sleep in PD, including exercise and physical activity, which have been shown promising results in improving sleep quality(Amara and Memon, 2018. Interestingly, studies have demonstrated an independent relationship between reduced sleep and sedentary behavior in PD (29). It has been suggested that treating sleep deficits may lead to a decline in sedentary behavior in the early stages of PD (5). This indicates the potential bidirectional relationship between sleep and sedentary behavior in PD. Although research on this topic is limited, it has been demonstrated that an independent and unique link exists between sedentary behavior and poor sleep quality(29).

### 4.4 Limitations

It is important to note that the evidence provided in this paper is not without limitations. A meta-analysis was not performed due to the heterogeneity of methods across the studies. Furthermore, studies included in this paper were collected from peer-reviewed journals via electronic databases. Thus, studies selected for this paper could potentially be subject to publication bias. Finally, data extraction and quality assessment were performed by one reviewer, hence the possibility of conceptual bias can not be excluded.

## 5. Future Goals and Directions

It is now well-established that non-motor symptoms of PD are a major contributor to the reduced quality of life in individuals living with PD(3,39). Despite often preceding the motor symptoms, non-motor symptoms have not received sufficient attention and remain undertreated (48). Currently, various pharmacological as well as non-pharmacological treatments are being investigated for both motor and non-motor symptoms of PD (18). Exercise has emerged as a promising non-pharmacological approach for improving not only motor symptoms but also non-motor symptoms(14). However, it is essential to recognize that sedentary behavior is an independent risk factor for poor health outcomes (16). Individuals with PD often lead sedentary lifestyles, making it crucial to assess its specific role in the pathophysiology of non-motor symptoms in PD in more depth. (13)Emerging research has highlighted the unique and crucial role of sedentary behavior in PD(13). For example, it has been demonstrated that individuals with PD tend to have longer bouts of sedentary behavior compared to their age-matched controls, suggesting a clear change in their pattern of sedentary behavior (49).

Despite its potential significance, current data on the role of sedentary behaviour in PD, particularly in the context of non-motor symptoms, is severely limited. Only 7 studies were found relevant to our topic, indicating the need for further investigation in this area. A clearer understanding of the relationship between sedentary behavior and non-motor symptoms may help to establish better biomarkers of the disease at an earlier stage, leading to more effective therapies for PD.

Future studies using more objective device-based, larger sample sizes, clear definition of sedentary behavior, and improved study designs are warranted as they may provide a better and clearer insight into the consequences of sedentary behavior and its potential implications in the non-motor symptoms of PD. In conclusion, while exercise has shown promise in addressing motor and non-motor symptoms in PD, the specific impact of sedentary behavior on the disease remains an important area for further research. Understanding the role of sedentary behavior in PD may open new avenues for therapeutic interventions and lead to improved outcomes for individuals living with the disease.

## Data Availability

All relevant data are within the manuscript and its Supporting Information files

